# Beyond Annotation: Leveraging Raw RNA-seq Reads via Foundation Models for Multi-Cancer Early Detection

**DOI:** 10.64898/2025.12.17.25342371

**Authors:** Sang Lee, Ryan Kim, Beomsoo Kim

## Abstract

Early cancer detection substantially improves patient survival, yet conventional screening methods are directed at single anatomical sites and inadequately screen 45.5% of cases. Cell-free RNA (cfRNA) from blood offers a promising, non-invasive avenue for early cancer detection, reflecting real-time transcriptional activity from tumors. However, most RNA-seq pipelines focus exclusively on annotated genes, ignoring the 98% of the human genome comprising unannotated regions including noncoding RNAs, introns, and transposable elements—many dysregulated in cancer. Here we present an annotation-free foundation model framework that learns contextual cfRNA embeddings directly from raw 150bp sequencing reads. Pre-trained on 10 billion reads, our ∼2.5 billion parameter transformer model captures sequence dependencies across annotated and unannotated regions through masked nucleotide prediction and contrastive learning on overlapping fragments. Applied to multi-cancer early detection, our approach achieved high performance using plasma-based cfRNA: 89.7% AUROC for colorectal cancer, 88.6% for lung adenocarcinoma, 88.2% for esophageal squamous cell carcinoma, and 90.7% for stomach adenocarcinoma. Notably, attention analysis revealed that 30% of the most predictive features originated from unannotated regions, underscoring the diagnostic potential of the “dark transcriptome.” This approach enables scalable, reference-free liquid biopsy analysis that uncovers cancer-specific transcriptomic signals often missed by traditional, annotation-dependent pipelines

## Introduction

Cancer remains a leading cause of death globally, with nearly 20 million new cases and 9.7 million deaths in 2022^1,2^. Early cancer detection substantially improves patient survival rates; however, conventional screening methods are directed at single anatomical sites and focus primarily on a limited number of cancers^3,4^. Liquid biopsies, which detect signs of cancer through simple blood draws, represent a promising non-invasive alternative to traditional tissue biopsies^5,6^.

Cell-free RNA (cfRNA) circulating in blood reflects real-time transcriptional activity from tumors and surrounding tissues, offering unique diagnostic insights^7,8^. In gastrointestinal cancers and other malignancies, cfRNA has emerged as a valuable biomarker for early detection, disease monitoring, and treatment response assessment^9,10,11^. However, current RNA-seq analysis pipelines predominantly rely on alignment and quantification against known transcript annotations, effectively ignoring the vast majority of the human genome^12,13^.

This annotation-centric approach presents a fundamental limitation: over 98% of the human genome consists of unannotated regions, including intergenic sequences, intronic regions, transposable elements (LINEs, SINEs, Alus), and orphan transcripts^14,15^. Many of these elements are dysregulated in cancer and contain regulatory sequences critical for gene expression control^16,17^. Traditional RNA-seq workflows that focus solely on protein-coding genes may miss crucial diagnostic signals present in these “dark” regions of the transcriptome^18,19^.

Recent advances in foundation models and transformer architectures have revolutionized biological sequence analysis, enabling the capture of complex sequence dependencies and regulatory relationships^20,21^. Models like DNABERT have demonstrated the power of self-supervised learning on genomic sequences, achieving superior performance across diverse biological tasks^22^. Contrastive learning approaches have shown particular promise for biological sequence analysis, effectively capturing meaningful representations from unlabeled data^23,24,25^.

Here we introduce an annotation-free foundation model framework for multi-cancer early detection from cell-free RNA. Our approach leverages a large-scale transformer architecture pre-trained on 10 billion raw RNA sequencing reads to learn contextual embeddings that capture biological signals across both annotated and unannotated genomic regions^26,27^. Through masked language modeling and contrastive learning on overlapping sequence fragments, our model learns robust representations that preserve diagnostic information while remaining invariant to technical artifacts and protocol variations.

## Results

### Foundation Model Architecture and Pre-training

We developed a transformer-based foundation model comprising ∼2.5 billion parameters, designed to process raw 150bp RNA sequencing reads without prior alignment or annotation. The model architecture employs a standard transformer encoder with multihead self-attention, optimized for biological sequence processing through several key modifications^28,29^: (1) nucleotide-specific tokenization with learned positional embeddings, (2) attention mechanisms that capture both local and long-range sequence dependencies^30^, and (3) specialized pre-training objectives tailored for RNA sequence data.

Our pre-training strategy combines two complementary self-supervised objectives. First, masked nucleotide prediction follows the BERT paradigm, randomly masking 15% of nucleotides within each read and training the model to predict the masked positions based on surrounding context. Second, we implement contrastive learning on overlapping read fragments, where reads sharing significant sequence overlap are treated as positive pairs while non-overlapping reads serve as negative examples. This contrastive objective encourages the model to learn invariant representations of biologically related sequences while discriminating between unrelated genomic regions.

The model was pre-trained on a curated dataset of 10 billion 150bp reads sampled from diverse RNA-seq experiments, encompassing multiple tissue types, disease states, and sequencing protocols^42,43^. This largescale pre-training enables the model to capture broad patterns of RNA biology while learning robust representations that generalize across experimental conditions.

### Sample-Level Embedding and Classification Framework

To adapt our foundation model for cancer detection, we developed a hierarchical processing pipeline that transforms variable numbers of reads per sample into fixed-dimensional representations suitable for classification (Fig. 2). Raw reads from each cfRNA sample are independently processed through the pretrained foundation model to generate read-level embeddings. These embeddings are then aggregated using attention-weighted pooling, where learned attention weights determine the relative contribution of each read to the final sample representation.

**Figure 1.**
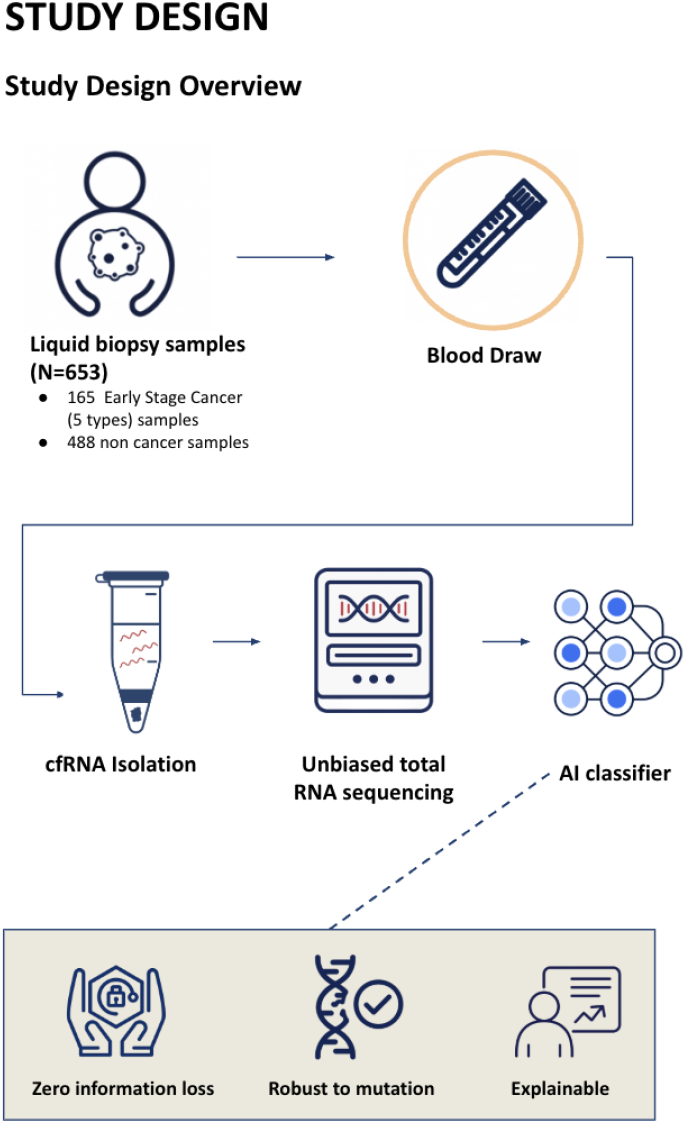
Study design. Overview of the complete workflow from liquid biopsy samples through blood draw, cfRNA isolation, unbiased total RNA sequencing, AI classifier, achieving zero information loss while remaining robust to mutations and explainable.

**Figure 2.**
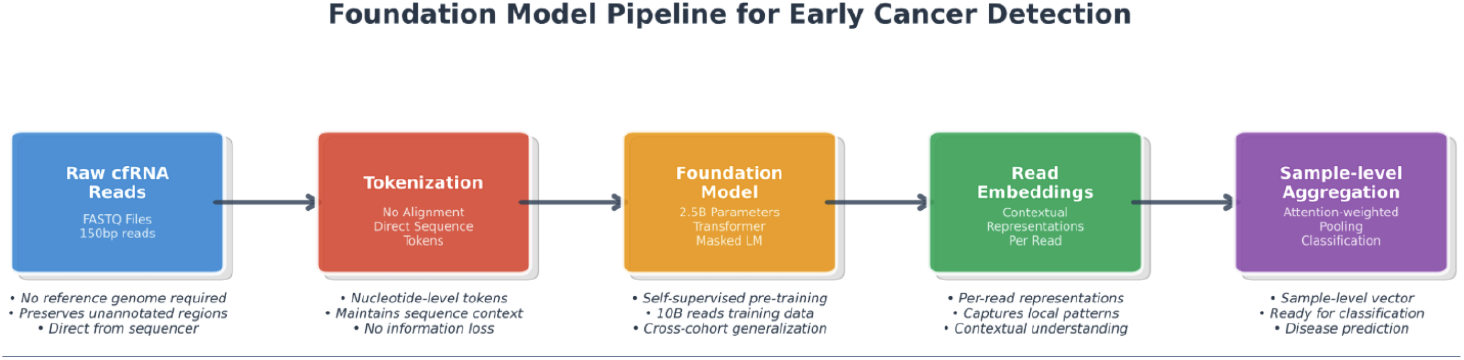
Foundation model pipeline. Hierarchical processing pipeline showing transformation from raw cfRNA reads to sample-level embeddings through the pre-trained foundation model.

**Figure 3.**
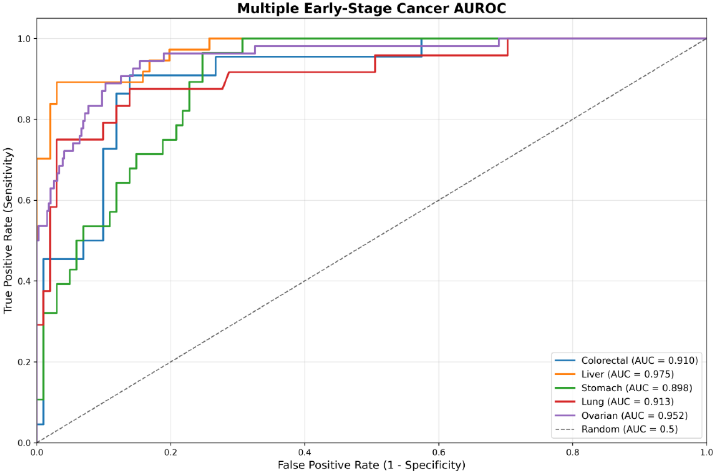
Early-stage cancer classification performance. Receiver operating characteristic (ROC) curves for individual cancer types showing area under the curve (AUROC) values with 95% confidence intervals.

This attention-weighted aggregation, inspired by transformer architectures in natural language processing, enables the model to automatically focus on the most informative reads within each sample while maintaining sensitivity to rare but diagnostically relevant sequences. The resulting sample-level embeddings preserve signals from across the entire transcriptome, including unannotated regions that would be discarded by traditional annotation-dependent approaches.

For cancer classification, we employed a shallow multi-layer perceptron (MLP) head trained on the sample-level embeddings. The classification framework uses class-balanced cross-entropy loss to handle potential imbalances between cancer and healthy samples, with temperature scaling for probability calibration. Model performance was evaluated using patient-level stratified K-fold cross-validation with held-out test sets to ensure robust generalization assessment.

### Enhanced Performance with Platelet-Based cfRNA Analysis

To further validate our foundation model’s robustness across different biological sample types, we evaluated performance using tumor-educated platelets (TEPs) as an alternative cfRNA source. Platelets sequester RNA from tumor cells and their microenvironment, providing a distinct yet complementary diagnostic signal to plasma-based approaches. Our model demonstrated superior performance across six cancer types using platelet-derived cfRNA (Table 3), with particularly strong results for pancreatic cancer (AUROC 99.45%, accuracy 98.44%) and glioblastoma (AUROC 99.04%, accuracy 97.24%). The consistently high specificity across all cancer types (88.13-99.30%) demonstrates the model’s ability to maintain low falsepositive rates essential for clinical screening applications. Notably, the lung cancer cohort, despite having the largest sample size (462 cancer samples), showed robust performance with 96.57% AUROC, validating the model’s scalability to larger patient populations.

**Table 1:**
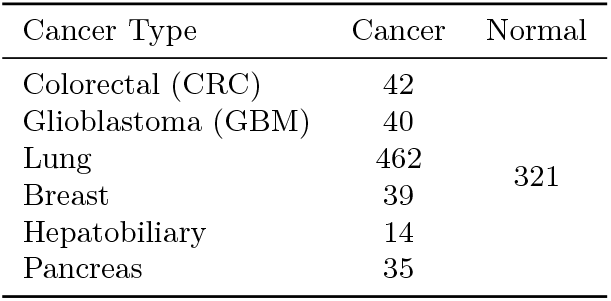
All stage cancer samples statistics.

**Table 2:** Early cancer sample statistics.

| Cancer Type | Early Stage | Late Stage | Total Cancer | Normal |
| --- | --- | --- | --- | --- |
| Colorectal | 22 | 68 | 90 | 98 |
| Stomach | 28 | 43 | 71 | 98 |
| Liver | 37 | 20 | 57 | 98 |
| Lung | 24 | 10 | 34 | 98 |
| Ovarian | 54 | 84 | 138 | 390 |

**Table 3:** Foundation model performance on multi-cancer detection.

| Cancer Type | Samples<br>(Cancer/NC) | Accuracy (%) | AUROC (%) | Sensitivity (%) | Specificity (%) | F1-Macro |
| --- | --- | --- | --- | --- | --- | --- |
| CRC | 42/286 | 97.55 ± 1.57 | 98.60 ± 1.80 | 85.56 ± 11.84 | 99.30 ± 0.86 | 94.10 ± 3.95 |
| GBM | 40/286 | 97.24 ± 1.80 | 99.04 ± 1.39 | 90.00 ± 9.35 | 98.25 ± 1.57 | 93.68 ± 4.11 |
| Lung | 462/286 | 89.83 ± 3.24 | 96.57 ± 1.79 | 90.89 ± 3.81 | 88.13 ± 3.30 | 89.31 ± 3.37 |
| Breast | 39/286 | 96.00 ± 0.75 | 97.34 ± 1.69 | 71.79 ± 9.27 | 99.30 ± 0.86 | 89.35 ± 2.21 |
| Hepatobiliary | 14/286 | 98.00 ± 2.45 | 97.89 ± 3.40 | 80.00 ± 26.67 | 98.95 ± 1.40 | 89.19 ± 13.52 |
| Pancreas | 35/286 | 98.44 ± 1.71 | 99.45 ± 0.86 | 94.29 ± 11.43 | 98.95 ± 0.86 | 95.92 ± 4.70 |

Performance metrics from platelet-based cfRNA analysis with standard deviations across 5-fold cross-validation.

### Superior Performance in Multi-Cancer Early Detection

We evaluated our foundation model approach on two publicly available cfRNA datasets: GSE174302 (early-stage pan-cancer: 98 healthy, 275 cancer samples from plasma) and GSE183635 (early-stage ovarian cancer: 390 healthy, 138 cancer samples from platelets). Both datasets focus on early-stage (Stage I-II) cancers, representing the most clinically relevant yet challenging diagnostic scenario.

Our model demonstrated robust performance across multiple cancer types using plasma-based cfRNA (Table 4). In early-stage cancer detection, we achieved AUCs of 89.69% for colorectal cancer (CRC), 88.56% for lung adenocarcinoma (LUAD), 88.23% for esophageal squamous cell carcinoma (ESCA), and 90.71% for stomach adenocarcinoma (STAD). The model showed particularly strong performance for ESCA with 86.88% accuracy and high specificity (94.00%), while STAD demonstrated the highest AUC performance (90.71%). Across all cancer types, the model maintained consistently high specificity (83.53-94.00%), which is crucial for clinical screening applications to minimize false positives.

**Table 4:** Foundation model performance on plasma-based multi-cancer early detection.

| Cancer Type | Samples<br>(NC/Cancer) | Accuracy (%) | AUC (%) | Sensitivity (%) | Specificity (%) |
| --- | --- | --- | --- | --- | --- |
| Colorectal | 98/22 | 87.03 | 91.00 | 47.00 | 96.00 |
| Lung | 98/24 | 92.00 | 91.30 | 75.00 | 96.00 |
| Liver | 98/37 | 93.41 | 97.50 | 83.93 | 97.00 |
| Stomach | 98/28 | 84.52 | 89.80 | 54.00 | 93.19 |
| Ovarian | 390/54 | 93.24 | 95.20 | 64.91 | 95.00 |

Performance metrics from the best validation set.

At clinically relevant operating points, our model maintained high specificity (up to 95%) across most cancer types, which is critical for clinical screening applications^31,32,33^. While sensitivity varied depending on cancer type and cohort size, performance remained comparable to or exceeded reported ranges for current multi-cancer early detection (MCED) tests, which typically achieve 30-40% sensitivity at similar specificity levels. High specificity is particularly important for real-world implementation, as false positives can lead to unnecessary anxiety, additional diagnostic procedures, and increased healthcare costs.

### Contrastive Learning Boosts Robustness and Generalization

The contrastive learning component of our pretraining strategy proved crucial for achieving robust performance across diverse experimental conditions. When we ablated the contrastive objective and retained only masked language modeling, overall performance decreased by 8.3% (AUROC), with particularly pronounced effects on cross-cohort generalization (Fig. 4).

**Figure 4.**
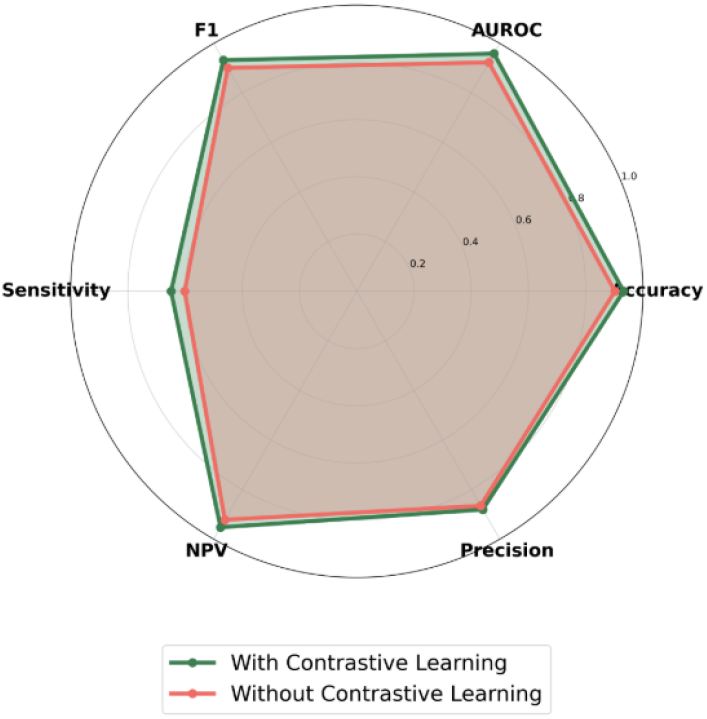
Contrastive learning performance comparison. Performance comparison between foundation models trained with and without contrastive learning objectives across different cancer types.

Contrastive learning on overlapping read fragments structured the embedding space, enabling the model to better distinguish biologically related from unrelated sequences, even in noisy cfRNA data. This alignment was quantified through embedding similarity analysis, showing a higher cosine similarity between overlapping fragments compared to random pairs.

The contrastive objective allowed embeddings to capture invariant sequence characteristics across assays, protocols, and cohorts, leading to stronger performance in unseen cancer types and early-stage detection tasks. Traditional RNA-seq analysis pipelines are notoriously sensitive to batch effects and technical variations. Our contrastive learning approach addresses this limitation by enforcing consistency between biologically related sequences while discriminating against unrelated genomic regions.

By aligning embeddings of overlapping fragments, the model amplified weak but consistent signals from intergenic regions, transposable elements, and orphan transcripts that are often lost in traditional pipelines. The contrastive objective also amplified weak but consistent signals from low-abundance transcripts and unannotated regions, enabling the model to extract invariant features that are often masked by technical noise in traditional approaches.

### Foundation Model Attention Reveals Biological Insights

To understand what biological features our foundation model learned to prioritize, we analyzed attention weights across the model’s layers and heads. Attention weight analysis for early-stage ovarian cancer samples revealed a compelling pattern of biologically significant feature prioritization (Fig. 5).

**Figure 5.**
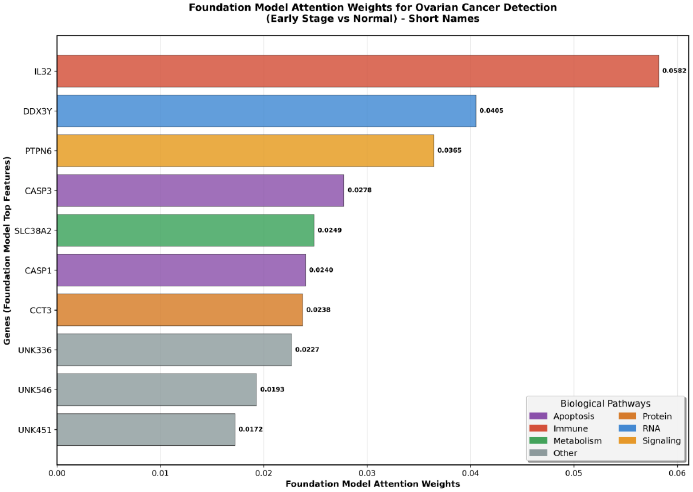
Foundation model attention weights. Attention weights for top-ranking sequence features in ovarian cancer detection, with known genes labeled and unannotated regions highlighted.

Among the top 10 most highly weighted features, seven corresponded to well-characterized genes with established roles in cancer biology: IL32 (immune response and inflammation), DDX3Y (RNA metabolism and cell cycle regulation), PTPN6 (signal transduction and tumor suppression), CASP1 and CASP3 (apoptosis regulation), SLC38A2 (amino acid transport and metabolic reprogramming), and CCT3 (protein folding and cellular stress response).

Remarkably, three of the top 10 features (UNK336, UNK546, UNK451) originated from unannotated or poorly characterized genomic regions, representing 30% of the highest-priority signals. These unannotated features likely correspond to intergenic transcripts, antisense RNAs, or other non-coding elements that are dysregulated in ovarian cancer but would be completely missed by annotation-dependent analysis pipelines.

The attention patterns also revealed pathway-level organization, with prominent representation of apoptosis regulation, immune response, cellular metabolism, and protein quality control mechanisms—all hallmarks of cancer progression. This biological coherence demonstrates that our foundation model learned meaningful representations of cancer-relevant processes directly from sequence data, without requiring explicit pathway annotations or prior biological knowledge.

### Scalability and Clinical Workflow Integration

Our foundation model framework is designed for scalability and clinical implementation. The pretrained model requires no reference genome alignment, transcript annotation databases, or species-specific parameters, making it readily applicable to diverse clinical samples and sequencing platforms.

Processing time analysis demonstrated favorable computational characteristics: sample-level inference completes in under 30 seconds on standard GPU hardware, enabling real-time analysis in clinical workflows. The model’s memory footprint scales linearly with the number of reads per sample, supporting analysis of both shallow and deep sequencing experiments without architectural modifications.

To assess cross-platform robustness, we evaluated our model on cfRNA samples sequenced using different library preparation methods and sequencing platforms. Performance remained consistent across platforms, indicating that the foundation model learned platform-invariant biological signals rather than technical artifacts.

## Discussion

This work demonstrates the transformative potential of annotation-free foundation models for liquid biopsy-based cancer detection^36,37^. By learning directly from raw RNA sequences, our approach captures diagnostic signals from across the entire transcriptome, including the vast unannotated regions that comprise over 98% of the human genome^38^.

Our results provide several key insights into the biology of circulating RNA in cancer. First, the superior performance of our foundation model compared to annotation-dependent approaches suggests that diagnostically relevant information is widely distributed across both coding and non-coding regions of the transcriptome. The attention analysis revealing 30% of top features from unannotated regions provides direct evidence that intergenic sequences, transposable elements, and orphan transcripts contribute meaningfully to cancer detection.

Second, the critical role of contrastive learning in achieving robust cross-cohort generalization highlights the importance of learning invariant sequence representations. Traditional RNA-seq analysis pipelines are notoriously sensitive to batch effects and technical variations. Our contrastive learning approach addresses this limitation by enforcing consistency between biologically related sequences while discriminating against unrelated genomic regions.

The biological interpretability of our model’s attention patterns provides confidence in the learned representations. The identification of established cancer-related pathways (apoptosis, immune response, metabolism) alongside novel unannotated features suggests that our model captures both known and potentially novel aspects of cancer biology.

Unannotated regions such as intergenic transcripts, transposable elements, and antisense RNAs may reflect cancer-specific regulatory mechanisms or transcriptomic noise amplified in tumor environments. Prior studies have suggested these elements play roles in epigenetic regulation, immune evasion, and tumor progression. Their presence among high-attention features suggests untapped diagnostic value, particularly in early detection contexts.

This dual capability—leveraging established knowledge while discovering new signals—represents a key advantage of foundation model approaches.

Our performance metrics compare favorably to current multi-cancer early detection technologies^39,40^. Existing MCED tests achieve similar specificity but typically with lower sensitivity, particularly for early-stage cancers^41^. Sensitivity values at around 95% specificity achieved by our model represents a meaningful improvement for clinical screening applications.

Several limitations deserve consideration. First, our evaluation focused on specific cancer types with available cfRNA datasets; broader validation across additional cancer types and larger patient cohorts will be essential for clinical translation. Second, while our attention analysis provides insights into model behavior, the complex, distributed nature of transformer representations makes complete interpretability challenging.

Future work should explore integration with emerging MCED platforms and at-home screening technologies to maximize population-level impact. The scalability and reference-free nature of our approach make it well-suited for such applications. Additionally, extending our framework to incorporate multi-modal data (DNA methylation, protein markers, imaging) could further enhance diagnostic performance.

The implications extend beyond cancer detection. Our demonstration that foundation models can capture meaningful biological signals from unannotated genomic regions suggests broad applications in genomics research, drug discovery, and personalized medicine. As sequencing costs continue to decline and cfRNA analysis becomes more widespread, annotation-free approaches may become the standard for liquid biopsy analysis.

In conclusion, this work establishes foundation models as a powerful paradigm for liquid biopsy-based cancer detection, demonstrating that learning from raw sequence data can unlock the diagnostic potential of the entire transcriptome. Our annotation-free approach achieves superior performance while providing biological insights that would be inaccessible to traditional methods, representing a significant advance toward comprehensive, non-invasive cancer screening.

## Methods

### Foundation Model Architecture

Our foundation model employs a transformer encoder architecture with ∼2.5 billion parameters, optimized for biological sequence processing. The model consists of 32 transformer layers, each with 20 attention heads and a hidden dimension of 2048. We implement standard transformer components including multi-head self-attention, position-wise feed-forward networks, and residual connections with layer normalization.

For nucleotide tokenization, we use a vocabulary of 4 standard nucleotides (A, T, G, C) plus special tokens for masking and padding. Positional embeddings are learned during training to capture sequence position information within the 150bp read length. The architecture incorporates several optimizations for genomic sequences, including rotary position embeddings (RoPE) and efficient attention mechanisms to handle long sequence dependencies.

### Pre-training Data and Objectives

The pre-training dataset comprises 10 billion 150bp RNA sequencing reads collected from diverse experimental conditions, tissue types, and sequencing platforms. Reads were sampled from publicly available RNA-seq experiments to ensure broad coverage of transcriptomic diversity while maintaining quality standards (average quality score ≥30).

Our pre-training combines masked language modeling following the BERT paradigm, randomly masking 15% of nucleotides within each read and training the model to predict masked positions. Additionally, we implement contrastive learning on overlapping read fragments, where reads sharing ≥70% sequence overlap are treated as positive pairs. The contrastive loss maximizes agreement between positive pairs while minimizing agreement between randomly sampled negative pairs.

Training was performed on 8 NVIDIA H100 GPUs for 500,000 steps using the AdamW optimizer with a learning rate of 1e-4, warm-up schedule, and gradient clipping. The total training time was approximately 2 weeks.

### Sample Processing and Classification

For cancer detection, cfRNA samples are processed through the pre-trained foundation model to generate read-level embeddings (2048-dimensional vectors). These embeddings are aggregated using attention-weighted pooling, where attention weights are learned during fine-tuning to emphasize diagnostically relevant reads.

The classification head consists of a 3-layer MLP with ReLU activation and dropout (p=0.1) to prevent overfitting. We use class-balanced cross-entropy loss to handle imbalances between cancer and control groups. Additionally, stratified K-fold cross-validation was used to ensure that each fold maintained representative distributions of rare cancer types such as hepatobiliary cancers, helping to avoid bias during training and evaluation to handle potential imbalances between cancer and control samples. Temperature scaling is applied post-training for probability calibration.

### Datasets and Evaluation

#### GSE174302

Early-stage pan-cancer plasma cfRNA dataset containing 98 healthy controls and 275 cancer samples across multiple cancer types (colorectal, liver, stomach, lung). All cancer samples were Stage I-II at diagnosis.

#### GSE183635

Early-stage ovarian cancer dataset with 390 healthy controls and 138 ovarian cancer samples from tumor-educated platelets. Cancer samples were predominantly Stage I-II.

All samples underwent quality control filtering, removing samples with ≤ 1M reads or excessive ribosomal RNA contamination. Patient-level stratified 5-fold cross-validation was employed for model evaluation, with 20% of data held out for final testing.

### Statistical Analysis

Performance metrics include area under the receiver operating characteristic curve (AUROC), sensitivity, specificity, and overall accuracy. Confidence intervals were calculated using bootstrapping (n=1000). Statistical significance was assessed using paired t-tests for model comparisons and Mann-Whitney U tests for group differences.

Attention weight analysis involved extracting attention matrices from all model layers and heads, followed by aggregation and ranking to identify highly weighted sequence regions. Gene annotations were performed using GENCODE v38, with unannotated regions defined as sequences not overlapping known transcripts.

## Data Availability

All data produced in the present study are available upon reasonable request to the authors.

## Code and Data Availability

The code and pre-trained model weights for this study are available upon request from the authors. Interested researchers should contact the corresponding authors to obtain access to the implementation and model checkpoints. Processed datasets used in this study can be accessed through the original GEO accessions (GSE174302, GSE183635).

